# Healthcare resource utilization and costs associated with COVID-19 among pediatrics managed in the community or hospital setting in England: a population-based cohort study

**DOI:** 10.1101/2023.07.28.23293335

**Authors:** Jingyan Yang, Kathleen M. Andersen, Kiran K. Rai, Theo Tritton, Tendai Mugwagwa, Carmen Tsang, Maya Reimbaeva, Leah J. McGrath, Poppy Payne, Bethany Backhouse, Diana Mendes, Rebecca Butfield, Robert Wood, Jennifer L. Nguyen

## Abstract

**Background:** Although COVID-19 morbidity is significantly lower in pediatrics than in adults, the risk of severe COVID-19 may still pose substantial healthcare resource burden. This study aimed to describe healthcare resource utilization (HCRU) and costs associated with COVID-19 in pediatrics aged 1-17 years in England.

**Methods:** A population-based retrospective cohort study of pediatrics with COVID-19 using Clinical Practice Research Datalink (CPRD Aurum) primary care data and, where available, linked Hospital Episode Statistics Admitted Patient Care (HES APC) secondary care data. HCRU and associated costs to the National Health Service (NHS) were stratified by age, risk of severe COVID-19, and immunocompromized status, separately for those with and without hospitalization records (hospitalized cohort: COVID-19 diagnosis August 2020-March 2021; primary care cohort: COVID-19 diagnosis August 2020-January 2022).

**Results:** This study included 564,644 patients in the primary care cohort and 60 in the hospitalized cohort. Primary care consultations were more common in those aged 1-4 years (face-to-face: 4.3%; telephone: 6.0%) compared to those aged 5-11 (2.0%; 2.1%) and 12-17 years (2.2%; 2.5%). In the hospitalized cohort, mean [SD] length of stay was longer (5.0 [5.8] days) among those aged 12-17 years (n=24) than those aged 1-4 (n=15; 1.8 [0.9] days) and 5-11 years (n=21; 2.8 [2.1] days).

**Conclusions:** Most pediatrics diagnosed with COVID-19 were managed in the community. However, hospitalizations were an important driver of HCRU and costs, particularly for those aged 12-17 years. Our results may help optimize the management and resource allocation of COVID-19 in this population.

## INTRODUCTION

Pediatrics with COVID-19 typically present asymptomatically or with mild symptoms.^1^ Hospitalization for COVID-19 and death are more rare in pediatrics compared to adults.^2, 3^ Yet, when pediatrics are hospitalized with COVID-19, 8-22% ^4–7^ require critical care according to United Kingdom (UK) and international estimates, and 4-9% require respiratory support.^6, 8^ Underlying health conditions ^8–10^ and multisystem inflammatory syndrome (MIS-C)^8^ are important risk factors for severe COVID-19 in pediatrics, and therefore the UK offered COVID-19 vaccines to all pediatrics aged ≥12 years by October 2021,^11^ which was extended to ages 5-11 years in April 2022.^12^ In April 2023 published guidance advised pediatrics aged 6 months to 4 years who are in a clinical risk group should be offered a COVID-19 vaccine.^13^ However, there remains a gap in understanding of what the economic burden of COVID-19 among pediatrics is, and therefore further research is needed to identify potential target populations for healthcare budget allocation and ultimately inform health policy decisions.

There is limited data on pediatric healthcare resource utilization (HCRU) following COVID-19 diagnosis ^4, 5, 14, 15^; these studies were either: 1) predominantly based on United States (US) data, therefore have limited generalisability to the UK, 2) lacked full coverage of childhood ages, and 3) did not report COVID-19-related primary care resource use e.g. general practitioner (GP) consultations. Similarly to the experience of hospitalizations, use of COVID-19 primary care services is likely to vary depending on age and presence of underlying health conditions; however age and comorbidity specific variations in GP or nurse visits have not been studied in pediatrics with COVID-19.

### Aims and objectives

To quantify HCRU and direct costs associated with COVID-19 in pediatrics to the National Health Service (NHS) in England, by age, risk of severe COVID-19 disease, and immunocompromized status, separately for those with and without hospitalization records, using primary care data linked to secondary care data where available.

## MATERIALS AND METHODS

### Study design and setting

We conducted a population-based retrospective cohort study using primary care data from the Clinical Practice Research Datalink (CPRD-Aurum)^16^ and linked secondary care administrative data from Hospital Episode Statistics Admitted Patient Care dataset (HES APC) where available.^17^ The May 2022 release of CPRD Aurum was used, which covers the period January 1995 to April 2022,^18, 19^ while the latest HES APC release covers April 1997 to March 2021.^17^ The study design and methods have been described elsewhere.^20^ A study design schematic is displayed in **Figure 1**.

**Figure 1.**
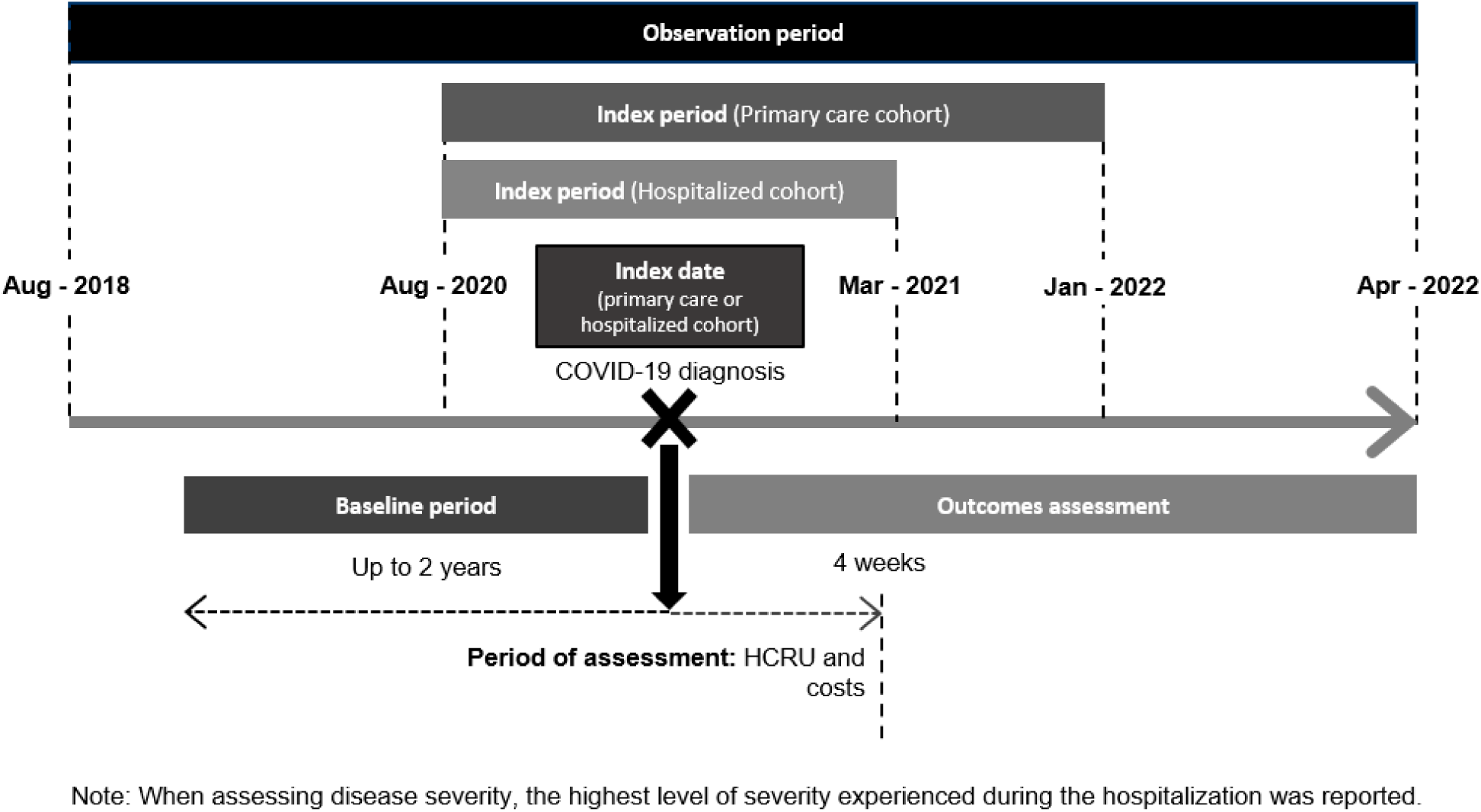
Study design schematic.

Two distinct patient cohorts were created to describe the economic burden of the acute phase (days 0-27 after COVID-19 diagnosis) among pediatrics in England:

1. **Hospitalized cohort:** Patients who had a positive SARS-CoV-2 polymerase chain reaction (PCR) or antigen test, or a recorded clinical diagnosis of COVID-19, in their GP record between August 1, 2020 and March 31, 2021 and had a hospitalization with a primary diagnosis of COVID-19 (ICD-10 U07.1 “COVID-19”) within 84 days after their GP recorded diagnosis. Details on the chosen index period and 84 days as a definition of hospitalization related to prior diagnosis are described elsewhere.^22^ Patients in this cohort may have also received COVID-19 care outside of the hospital setting, (e.g. primary care consultations).
2. **Primary care cohort:** Persons diagnosed with COVID-19 between August 1, 2020 and March 31, 2021 that did not have a record for a COVID-19 related hospitalization within 84 days of their positive test result *as well as* all persons diagnosed on or after April 1, 2021 (the period of time for which CPRD did not have hospitalization data available) were included in this cohort. The index period end date, January 2022, was chosen, allowing for adequate follow-up time.^20^

### Population

Patients aged 1-17 years were included in this study, with eligibility criteria cited elsewhere.^20^

### Demographic and clinical characteristics

Sociodemographic characteristics at index included: age, sex, region of GP practice, ethnicity and social deprivation (measured using quintiles of the 2019 Index of Multiple Deprivation (IMD) score). Clinical characteristics included Quan-Charlson Comorbidity Index (CCI) 2005^21^ within two years prior to index. For immune system status, patients were classified as immunocompromized at the time of receipt of first COVID-19 vaccine dose.^22^ Vaccination status was defined according to the number of primary series and booster doses received ≥14 days prior to COVID-19 diagnosis.^20^ Disease severity among the hospitalized cohort was assessed using the Ordinal Scale for Clinical Improvement within the World Health Organisation’s (WHO’s) COVID-19 Therapeutic Trial Synopsis,^23^ based on the highest level of care received during the hospitalization: 1) hospitalized, no oxygen therapy; 2) oxygen by mask or nasal prongs; 3) non-invasive ventilation or high-flow oxygen; 4) intubation and mechanical ventilation and 5) ventilation and additional organ support. Further details on definitions, and operationalization of code lists are described elsewhere.^20^

### Outcomes and follow-up

#### HCRU

All COVID-19 related HCRU and associated costs to the NHS in the four weeks including and following the index date were calculated and reported for the following elements:

**Medication use:** A pre-defined list of medications for the treatment of COVID-19 that were prescribed within primary care were considered to be COVID-19 related when prescribed on the same day as a COVID-19 diagnosis and included: nirmatrelvir/ritonavir, 2) remdesivir, 3) sotrovimab, 4) molnupiravir, 5) dexamethasone, 6) tocilizumab, 7) sarilumab, 8) casirivimab/imdevimab and 9) baricitinib.^24, 25^

**Primary care consultations:** GP or nurse consultations with a diagnostic code of COVID-19 were reported separately for face-to-face (F2F) and telephone consultations. This was defined as a maximum of one visit of each format per person per day, and any additional visits were considered as data capture errors.

**Hospitalizations:** Length of stay (LoS) per admission was reported separately for the overall admission as well as time spent in high dependency/intensive care units (HDU/ICU). In the event of multiple hospitalizations (or for critical care, HDU/ICU stays) during the acute COVID-19 phase, the average LoS per person, rather than the cumulative total, was used.

**Direct healthcare costs:** Costs were described for patients with ≥1 event of a given type only, (i.e. resource users); persons without utilization were not included in the distributions of costs presented. Hospitalization-associated costs were estimated based on the National Schedule of NHS Costs (2020/2021), which report costs of admitted patient care by Healthcare Resource Group (HRG) in England.^26^ HRGs were derived using the HRG4+ 2021/2022 National Costs Grouper distributed by the NHS.^27^ In order to estimate the cost per hospitalization, all finished consultant episodes (FCEs: the time a patient spends in the care of one consultant within their hospitalization) within each admission were accounted to derive the total spell (hospitalization) cost.^28^ Given patients could have multiple hospitalizations during the acute phase, cost for all hospitalizations, per patient, were reported.

Primary care consultations (including GP and nurse visits) were costed using information by the Personal Social Services Research Unit (PSSRU).^29^ The direct healthcare cost for each prescription written in primary care was calculated via the application of list price published in NICE guidance consultation for therapeutics for people with COVID-19 (version November 2022).^30^

### Statistical analysis

Means (SD), or medians (Q1, Q3) were calculated for numeric variables, with frequency counts and percentages presented for categorical variables. Results are presented separately for the hospitalized cohort and primary care cohort. Data on <5 patients were suppressed to comply with CPRD reporting rules, with secondary suppression implemented where relevant.

### Stratifying variables

Outcomes were evaluated by age group, high risk status, and immunocompromized status (primary care cohort only), as healthcare utilization can differ by age and clinical status.^31, 32^ Age categories were based on the COVID-19 vaccination rollout strategy in the UK: 1-4, 5-11, and 12-17 years. The UK’s COVID-19 vaccination prioritization criteria, as outlined in the Green Book chapter 14a, was used to define persons at greater risk of severe COVID-19.^20, 33^ All analyses were conducted in SAS version 9.4 (SAS Institute, Cary, North Carolina).

## RESULTS

### Patient sociodemographic and clinical characteristics

A total of 564,704 COVID-19 cases were included in this study. Among patients diagnosed in the time period for which hospitalization data were available, 0.1% (n=60 out of 64,325 cases through March 31, 2021) were hospitalized. In addition to the 64,265 non-hospitalized cases, there were also 501,399 patients diagnosed in the period after hospitalization data ended, together forming a primary care cohort of 564,644 people. **Table 1** summarizes the baseline patient characteristics across the primary care and hospitalized cohorts.

**Table 1:** Sociodemographic and clinical characteristics of the study population at baseline.

| Characteristics | Primary care cohort<br>(n=564,644)<br>n (%) | Hospitalized cohort<br>(n=60)<br>n (%) |
| --- | --- | --- |
| **Age, years** |  |  |
| Mean (SD) | 10.8 (4.0) | 9.7 (5.3) |
| 1-4 | 40,658 (7.2%) | 15 (25.0%) |
| 5-11 | 260,186 (46.1%) | 21 (35.0%) |
| 12-17 | 263,800 (46.7%) | 24 (40.0%) |
| **Sex** |  |  |
| Male | 281,588 (49.9%) | 33 (55.0%) |
| Female | 283,046 (50.1%) | 27 (45.0%) |
| Unknown | 10 (<0.1%) | 0 |
| **GP practice region** |  |  |
| North East | 18,079 (3.2%) | <5 |
| North West | 107,425 (19.0%) | 14 (23.3%) |
| Yorkshire and The Humber | 17,652 (3.1%) | <5 |
| East Midlands | 10,988 (2.0%) | <5 |
| West Midlands | 98,955 (17.5%) | 8 (13.3%) |
| East of England | 28,693 (5.1%) | <5 |
| London | 89,675 (15.9%) | 19 (31.7%) |
| South East | 130,638 (23.1%) | 12 (20.0%) |
| South West | 62,468 (11.1%) | <5 |
| Unknown | 71 (<0.1%) | 0 |
| **Ethnicity** |  |  |
| White | 413,741 (73.3%) | 36 (60.0%) |
| Black | 13,440 (2.4%) | 9 (15.0%) |
| Asian | 42,134 (7.5%) | 5 (8.3%) |
| Mixed | 19,294 (3.4%) | <5 |
| Other | 11,717 (2.1%) | <5 |
| Unknown | 64,318 (11.4%) | 6 (10.0%) |
| **Index of Multiple Deprivation (2019)** |  |  |
| Quintile 1 (least deprived) | 136,910 (24.3%) | 8 (13.3%) |
| Quintile 2 | 116,404 (20.6%) | 7 (11.7%) |
| Quintile 3 | 104,127 (18.4%) | 9 (15.0%) |
| Quintile 4 | 101,580 (18.0%) | 18 (30.0%) |
| Quintile 5 (most deprived) | 105,337 (18.7%) | 18 (30.0%) |
| Unknown | 286 (0.1%) | 0 |
| **Quan-CCI (2005)** |  |  |
| Mean (SD) | 0.1 (0.2) | 0.2 (0.6) |
| 0 | 540,466 (95.7%) | 50 (83.3%) |
| 1-2 | 23,813 (4.2%) | NR |
| 3-4 | 314 (0.1%) | <5 |
| ≥5 | 51 (<0.1%) | 0 |
| **Immunocompromized status** |  |  |
| Immunocompetent | 563,525 (99.8%) | 60 (100%) |
| Immunocompromized | 1,119 (0.2%) | 0 |
| **Vaccination status: immunocompetent** |  |  |
| Unvaccinated | 509,899 (90.5%) | 60 (100%) |
| 1 dose | 48,724 (8.7%) | 0 |
| 2 doses | 4,810 (0.9%) | 0 |
| First booster dose | 92 (<0.1%) | 0 |
| **Vaccination status: immunocompromized** |  |  |
| Unvaccinated | 152 (13.6%) | 0 |
| 1 dose | 719 (64.3%) | 0 |
| 2 doses | 222 (19.8%) | 0 |
| 3 doses | 26 (2.3%) | 0 |
| First booster dose | 0 | 0 |
| **Risk of severe COVID-19** |  |  |
| Defined per the Green Book | 12,495 (2.2%) | 12 (20.0%) |
| **COVID-19 severity<sup>a</sup>** | - |  |
| No oxygen therapy | - | 56 (93.3%) |
| Low-flow oxygen | - | <5 |
| Non-invasive ventilation or high-flow oxygen | - | 0 |
| Intubation/mechanical ventilation | - | <5 |
| Ventilation and additional organ support | - | 0 |

<sup>a</sup> Highest level of severity experienced during the hospitalization, not measured within baseline.
NR: Not reported due to secondary suppression to comply with CPRD reporting rules.
Note, the hospitalized cohort included patients with documented COVID-19 in their GP records between 1 August 2020 and 31 March 2021 and had a COVID-19 hospitalization within 84 days of index. Patients with documented COVID-19 in their GP records between 1 August 2020 and 31 March 2021 that did not have a record for a COVID-19 related hospitalization within 84 days of index *as well as* all persons diagnosed on or after 1 April 2021 (the period of time for which CPRD did not have hospitalization data available) were included in this cohort.

#### Primary care cohort

Similar proportions were observed in those aged 5-11 (46.1%) and 12-17 years (46.7%), with 7.2% in the 1-4 age group (mean age [SD]: 10.8 [4.0]). Half were female (50.1%), the majority were of white ethnicity (73.3%), and 36.7% lived in areas of higher deprivation (2019 IMD quintiles 4 and 5). Most (95.7%) had a CCI score of 0, and among those immunocompetent at baseline (99.8%), 90.5% were unvaccinated at index. Only 2.2% were at high risk of severe disease.

#### Hospitalized cohort

Baseline sociodemographic characteristics in the hospitalized cohort (n=60) differed numerically to the primary care cohort. A greater proportion in the hospitalized cohort than in primary care cohort were aged 1-4 years (25.0% vs. 7.2%), while fewer were aged 5-11 years (35.0% vs. 46.1%) and 12-17 years (40.0% vs. 46.7%). Hospitalized patients were more often male (55.0% vs. 49.9%), less often white ethnicity (60.0% vs 73.3%), more often lived in areas of higher deprivation (60.0% vs 36.7%) and were more often at high risk (20.0% vs. 2.2%) than those in the primary care cohort. All were considered immunocompetent and unvaccinated at index. Most (93.3%) did not receive any oxygen therapy or respiratory support.

### HCRU and associated costs of the acute phase of COVID-19

#### Primary care cohort

A total of 24,582 (4.6%) of pediatrics had a primary care consultation (with either a GP or nurse). Similar proportions of patients in the overall group experienced at least one F2F consultation (2.3%) and at least one telephone consultation (2.6%) **(Table 2)**. The mean cost of F2F consultations was highest among those aged 1-4 years (mean: £31 [SD: 17]) and similar across the other stratifications (overall: £26 [18]; 5-11 years: £25 [17]; 12-17 years: £25 [17]; high risk : £28 [18]; immunocompetent: £26 [18]), except among the immunocompromized group (£20 [19]).

**Table 2:**
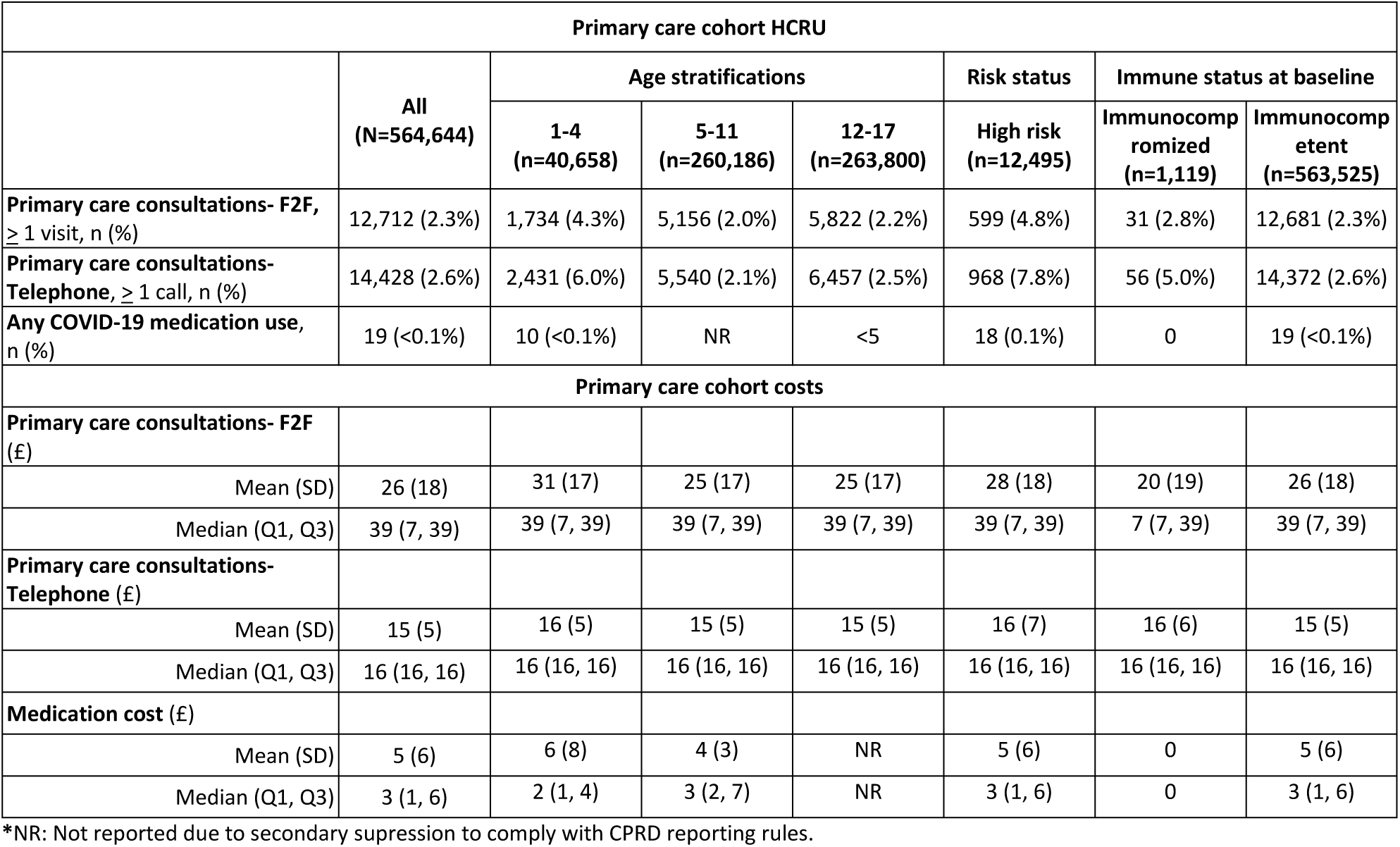
HCRU and costs in the acute phase of COVID-19 stratified by age, risk of severe COVID-19 and immune status at baseline among the primary care cohort.

Primary care telephone consultations were more common in pediatrics aged 1-4 years (6.0%), among high risk (7.8%), and immunocompromized pediatrics (5.0%). For telephone consultations, the overall mean cost was £15 [5], and was similar across all stratifications.

Overall, the frequency of prescriptions for COVID-19-related medication in primary care was low (n=19; <0.1%; all patients receiving these prescriptions were immunocompetent, and the majority (n=18) were at high risk.

#### Hospitalized cohort

The mean total LoS within the acute phase was 3.4 days (SD: 4.0), and was highest among patients aged 12-17 years (mean: 5.0 days [SD: 5.8]) compared to 1-4 years (1.8 days [0.9]), 5-11 years (2.8 days [2.1]) and high risk patients (3.2 days [3.8]) **(Table 3)**. Five patients (8.3%) were admitted to critical care and among these the mean LoS within a critical care admission was 4.0 days (SD: 1.9). There were no patients in the 1-4 or 5-11 year groups with a readmission in the acute phase, and <5 patients aged 12-17 years with a readmission in the acute phase.

**Table 3:**
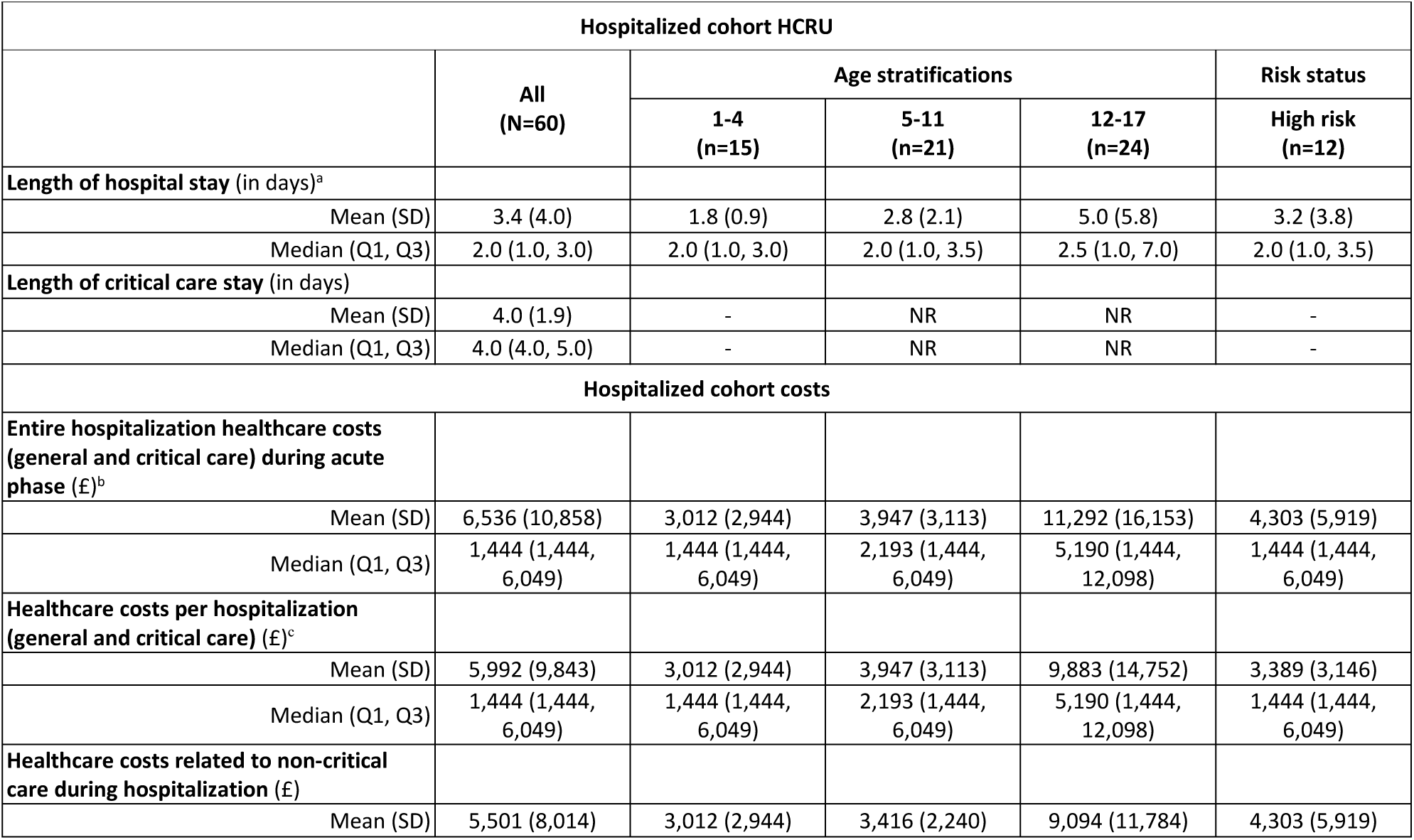
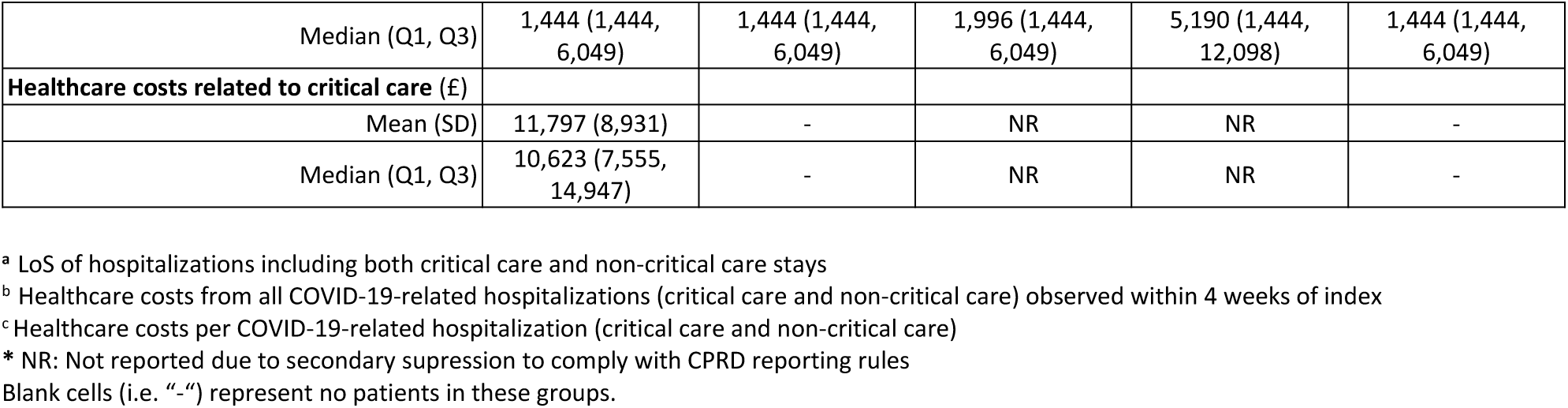
HCRU and costs in the acute phase of COVID-19 stratified by age and risk of severe COVID-19 at baseline among the hospitalized cohort.

Mean cost per patient for all hospitalizations within 4 weeks of index was higher (mean £6,536 [10,858]) than the mean cost per hospitalization per patient (£5,992 [9,843]), where both estimates included general and critical care costs. Cost per hospitalization was higher among patients aged 12-17 years (£9,883 [£14,752]) compared to younger age groups (1-4 years: £3,012 [2,944]; 5-11 years: 3,947 [3,113]). Similar trends were observed for costs associated with non-critical care (1-4 years: £3,012 [2,944]; 5-11 years: £3,416 [2,240]; 12-17 years: £9,094 [11,784]). Overall mean critical care admission cost was £11,797 [SD: 8,931]. The mean number of FCEs per patient with a hospitalization in the acute phase was 1.3, with FCEs ranging between 1.0-5.0 (data not shown).

## DISCUSSION

To the best of our knowledge, this is the first study to quantify HCRU and related costs during the acute phase of COVID-19 among pediatrics aged 1-17 years in the primary and secondary care settings in England. We found primary care resource use and costs were greater among those aged 1-4 years and among those at risk of severe COVID-19 compared to those aged 5-17 years and the overall cohort, respectively. In comparison we observed greater secondary care resource use and costs in those aged 12-17 years compared to the younger age groups and the overall pediatric cohort. Our findings have highlighted an excess health care burden of COVID-19 in pediatrics, particularly among patients requiring hospitalization, which may provide valuable evidence to inform strategic resource allocation across care settings.

Our findings that patients aged 1-4 years more often had a primary care consultation than older pediatrics is consistent with prior studies using CPRD pre- and within-pandemic data.^34, 35^ In particular, Vestesson *et al*.^36^ found that between March 2018 and February 2022 patients aged <5 years sustained the highest consultation rates in the pediatric group regardless of the mode of consultation (remote versus F2F). These patterns may in-part reflect evidence showing that respiratory care for mild symptoms was less frequently sought for older pediatrics than younger pediatrics,^15^ and that younger pediatrics with respiratory infections are perceived as more vulnerable by their caregivers due to difficulties with communication and concerns of rapid deterioration of health in the younger ages.^37^

In the present study, the proportion of patients in the overall cohort with ≥1 F2F consultations was similar (2.3%) to the proportion with ≥1 telephone consultations (2.6%), which corresponds to Vestesson *et al*’s^36^ estimates during the pandemic. This may be partially explained by the rapid implementation of remote consultations in general practices in the UK due to the COVID-19 pandemic. ^38, 39^ Data from a longitudinal study demonstrated a drop in F2F consultations in all ages since the pandemic due to infection control, however this reduction was less for patients aged 0-4 years than aged 5-17 years.^34, 40^

We observed a higher proportion of primary care consultations in both pediatrics at higher risk of severe COVID-19 and in the immunocompromized group than in the overall cohort. We also observed proportionally greater healthcare resource use in high risk patients than in those who were immunocompromized, which is in keeping with literature that has shown that in pediatrics, being immunocompromized did not increase the risk of severe COVID-19,^41^ but that presence of underlying conditions is a key driver of increased risk of severe COVID-19.^9^

In the period for which HES APC data were available, the overall LoS in hospitalized pediatrics (3.4 days) was similar to the estimate within a US-based study using claims data (4.3 days).^5^ When stratified by age, the US study observed a longer LoS among those aged 0-4 years (4.1 days) and 5-11 years (4.7 days) compared to our study (LoS estimates including general and critical care: 1-4 years: 1.8 days; 5-11 years: 2.8 days). That said, trends showing increasing LoS with increasing age were comparable between studies. This shorter hospital duration in the younger pediatrics may be related to a risk averse culture, reduced access to primary care services, particularly out-of-hours, and parents conforming to social expectations to seek care when their child is sick.^42^ We identified 8.3% of pediatrics were admitted to critical care during our study period; this is higher than the national estimate of 4.1%,^43^ but our results should be interpreted with caution due to the small sample size.

Regarding the cost of a COVID-19 hospitalization, our data is similar to the estimated hospitalization costs in 2020 (British pound sterling, GBP) from a UK NHS perspective which demonstrates an average cost of £4847 per hospitalized patient, although this calculation included all ages.^44^

Our study has several limitations. First, our previous published work found an over-representation of patients living in certain English regions such as London and the South East.^20^ This may partly explain the lower proportion of younger pediatrics aged <5 years compared to the older ages in our hospitalized cohort; which is the reverse of the patterns observed from national and international estimates.^5, 45^ Due to HES APC data latency, hospitalization status was unknown for pediatrics in the primary care cohort after April 2021. Furthermore, the study observation period pre-dates omicron predominance, for which there is evidence that pediatrics were more susceptible but less frequently experienced severe disease and hospitalization following infection,^46, 47^ with UK data demonstrating the risk of hospitalization during omicron compared to delta was not significantly different in pediatrics <10 years.^48^

Lastly, as COVID-19 becomes endemic, with the associated removal of routine COVID-19 testing and national restrictions, COVID-19 related healthcare seeking behaviour in pediatrics may differ to that observed during the UK epidemic; therefore, HCRU and cost patterns in this study may not fully reflect the current and forthcoming period.

Our study showed that while most pediatrics with COVID-19 are managed in the community, hospitalization is an important driver of HCRU and costs, particularly for those aged 12-17 years. COVID-19 in pediatrics is associated with reduced health-related quality of life,^49^ school absence,^50^ increased family instability (such as disruption to family income and caregiver stressors), and onward transmission to other social contacts and family members.^51^ School absence in particular, associated with lower educational attainment^50^ and caregiver work absenteeism,^52^ poses a multi-faceted risk to pediatrics.

Future research with a larger sample size is required to better quantify the economic burden of COVID-19 among hospitalized patients, in particular, those who are immunocompromized and admitted to critical care. Studies should also explore the consequences of post-COVID conditions as well as the longer-term impact of COVID in relation to HCRU and related costs, school absence and the burden on caregivers.

### Conclusions

We observed pediatrics aged 1-4 years most frequently had a primary care interaction compared to other age groups. Despite a smaller number of cases requiring hospitalization, those who were admitted to hospital for COVID-19 contributed substantial economic burden. Findings from this study may help optimize the management and resource allocation of COVID-19 in pediatrics.

## Data Availability

All data produced in the present study are available upon reasonable request to the authors.

## Acknowledgments

The authors gratefully acknowledge Tamuno Alfred, Darren Kailung Jeng and Chern Chuan Soo from Pfizer Inc. (New York, United States), Agnieszka Gajewska, Tomasz Mikołajczyk, Ewa Śleszyńska-Dopiera from Quanticate (Warsaw, Poland) and Olivia Massey from Adelphi Real World for statistical programming support.

## Copyright

The corresponding author has the right to grant on behalf of all authors and does grant on behalf of all authors, a worldwide licence to the Publishers and its licensees in perpetuity, in all forms, formats and media (whether known now or created in the future), to i) publish, reproduce, distribute, display and store the Contribution, ii) translate the Contribution into other languages, create adaptations, reprints, include within collections and create summaries, extracts and/or, abstracts of the Contribution, iii) create any other derivative work(s) based on the Contribution, iv) to exploit all subsidiary rights in the Contribution, v) the inclusion of electronic links from the Contribution to third party material where-ever it may be located; and, vi) licence any third party to do any or all of the above.

## Author contributions

The corresponding author, Jingyan Yang, attests that all listed authors meet authorship criteria and that no others meeting the criteria have been omitted.

All authors were involved in: 1) conception or design, or analysis and interpretation of data; 2) drafting and revising the article; 3) providing intellectual content of critical importance to the work described; and 4) final approval of the version to be published, and therefore meet the criteria for authorship in accordance with the International Committee of Medical Journal Editors (ICMJE) guidelines. In addition, all named authors take responsibility for the integrity of the work as a whole and have given their approval for this version to be published.

## Ethical approval

CPRD’s Research Data Governance (RDG) committee approved this study (CPRD study ID: 22_002062) in July 2022 prior to obtaining the data relevant to the project. This study complied with all applicable laws regarding subject privacy. As all patient-level data were fully anonymised, and no direct patient contact or primary collection of individual patient data occurred, patient consent was not required.

## Dissemination to participants and related patient and public communities

The study findings will be disseminated to the public through appropriate channels.

## Data sharing statement

**Data may be obtained from a third party and are not publicly available.** Anonymised patient data were accessed under study-specific approvals. Electronic health records are considered sensitive data in the UK by the Data Protection Act and cannot be shared. Access to the primary care data and linked datasets could be requested from the Clinical Practice Research Datalink https://www.cprd.com/research-applications.

## Transparency declaration

The lead author affirms that this manuscript is an honest, accurate, and transparent account of the study being reported; that no important aspects of the study have been omitted; and that any discrepancies from the study as planned (and, if relevant, registered) have been explained.

## Prior publication

Data in this manuscript was accepted as one oral presentation at the European Society for Paediatric Infectious Diseases (ESPID) 2023, Lisbon, Portugal in May 2023 and two poster presentations to the International Society for Pharmacoeconomics and Outcomes Research (ISPOR) 2023, Boston, United States in May 2023.

